# Increased Expression of Colonic Mucosal Melatonin in Patients with Irritable Bowel Syndrome Correlated with Gut Dysbiosis

**DOI:** 10.1101/2020.03.03.20030635

**Authors:** Ben Wang, Shiwei Zhu, Zuojing Liu, Hui Wei, Lu Zhang, Fei Pei, Meibo He, Jindong Zhang, Qinghua Sun, Liping Duan

**Affiliations:** Department of Gastroenterology, Peking University Third Hospital, Beijing 100191, China; Department of Pathology, School of Basic Medical Sciences, Peking University, Beijing 100191, China; Institute of Systems Biomedicine, School of Basic Medical Sciences, Peking University, Beijing 100191, China

**Keywords:** Irritable bowel syndrome, Melatonin, Gut microbiota, Fecal microbiota transplantation, Butyrate

## Abstract

Dysregulation of the gut microbiota/gut hormone axis contributes to the pathogenesis of irritable bowel syndrome (IBS). Melatonin plays a beneficial role in gut motility and immunity. However, altered expression of local mucosal melatonin in IBS and its relationship with the gut microbiota remain unclear. Therefore, we aimed to detect the colonic melatonin levels and microbiota profiles in patients with diarrhea-predominant IBS (IBS-D) and explore their relationship in germ-free (GF) rats and BON-1 cells. Thirty-two IBS-D patients and twenty-eight healthy controls (HC) were recruited. Fecal specimens from IBS-D patients and HCs were separately transplanted into GF rats by gavage. The levels of colon mucosal melatonin were assessed by immunohistochemical methods, and fecal microbiota communities were analyzed using 16S rDNA sequencing. The effect of butyrate on melatonin synthesis in BON-1 cells was evaluated by ELISA. Melatonin levels were significantly increased in IBS-D patients compared with HC and were significantly negatively correlated with visceral sensitivity in IBS-D patients. GF rats inoculated with fecal microbiota from IBS-D patients had high colonic melatonin levels. Butyrate-producing *Clostridium* cluster XIVa species, such as *Roseburia* species and *Lachnospira* species, were positively related to colonic mucosal melatonin expression. Butyrate significantly increased melatonin secretion in BON-1 cells. Increased melatonin expression may be an adaptive protective mechanism in the development of IBS-D. Moreover, some *Clostridium* cluster XIVa species could increase melatonin expression via butyrate production. Modulation of the gut hormone/gut microbiota axis offers a promising target of interest for IBS in the future.

## Introduction

Irritable bowel syndrome (IBS) is a common functional gastrointestinal disease that considerably affects quality of life and work productivity [1]. Gut dysmotility, visceral hypersensitivity, intestinal barrier dysfunction, immune activation, altered microbiota, and brain-gut axis dysregulation may contribute to the development of IBS [2]. Additionally, mast cells have been suggested as noteworthy players in the pathophysiology of IBS [3]. Recently, gut hormones have attracted increasing attention in IBS development and aggravation. The secretion of cholecystokinin, glucagon-like peptide 1, peptide YY, and serotonin differs between patients with diarrhea-predominant IBS (IBS-D) and healthy controls (HC), which may contribute to the pathogenesis of IBS and the maintenance of symptoms [4].

Melatonin is widely found in nature [5,6]. In vertebrates, melatonin was first reported in the pineal gland. Subsequently, melatonin was also discovered in the retina, Harderian gland, and gastrointestinal tract (GIT). Enterochromaffin cells in the GIT mucosa have been shown to synthesize melatonin [7]. Accumulating evidence has suggested the important roles and activities of melatonin in the GIT, especially enteroprotective effects such as immunomodulatory activity, antioxidant effects, maintenance of gastric prostaglandin levels and promotion of bicarbonate secretion [8–10]. Additionally, melatonin has also been reported to be involved in the regulation of gut motility [11,12].

Given the importance of melatonin in gastrointestinal function, it is very likely that melatonin affects the pathophysiology of IBS. Some studies evaluated the effect of melatonin administration on IBS and found that it alleviates abdominal pain in IBS patients, but the mechanism remains unclear [13]. Therefore, exploration of the role of GIT melatonin in IBS is essential. However, the altered expression of local mucosal melatonin in IBS has not been evaluated to date.

Increasing evidence suggesting the effect of gut microbiota on the pathophysiology of IBS [14] and the importance of the gut microbiota/gut hormone axis in the pathogenesis of IBS has attracted increasing attention [4]. Additionally, a recent study suggested that melatonin could alleviate lipid metabolism in high fat diet-fed mice by modulating gut microbiota [15]. However, the association between mucosal melatonin and gut microbiota remains obscure.

The aims of our study were (1) to detect colonic mucosal melatonin levels and gut dysbiosis in patients with IBS-D and (2) to further assess whether altered melatonin expression correlates with gut microbiota in humans, GF rats and BON-1 cells (a model of enterochromaffin cells [16]).

## Results and discussion

### Demographic and clinical characteristics of patients with IBS-D

Thirty-two IBS-D patients and twenty-eight HC were included. The demographic and clinical characteristics are presented in **Table 1**. No significant differences in age, sex, or body mass index (BMI) were found between the two groups. Anxiety and depression scores were higher in patients with IBS-D, but the difference was not significant. Protein, fat and carbohydrate intake was described as a proportion of total energy consumption. No significant differences were found in the proportion of the energy intake from protein, fat or carbohydrates between the two groups (Figure S1). Twenty-eight IBS-D patients and seventeen HC underwent a rectal barostat test to evaluate visceral sensitivity, and the thresholds for sensation of defecation in IBS-D patients were significantly lower than those in HC (**Table 1)**.

**Table 1.** Demographic and clinical features of patients with IBS-D and HC.

|  | IBS-D | HC | *P* value |
| --- | --- | --- | --- |
| Number of subjects | 32 | 28 |  |
| Age (years) | 32 ± 1.77 | 30 ± 1.56 | 0.471 |
| Gender ( male:female) | 28:4 | 20:8 | 0.219 |
| BMI (kg/m2) | 22.6 ± 0.62 | 23.1 ± 0.73 | 0.564 |
| *Hospital anxiety scale | 5.41 ± 0.60 | 3.90 ± 0.79 | 0.127 |
| *Hospital depression scale | 4.28 ± 0.47 | 2.75 ± 0.65 | 0.056 |
| #Rectal distension (mmHg) |  |  |  |
| First sensation | 9.07 ± 0.60 | 11.14 ± 0.88 | 0.055 |
| Sensation of defecation | 16.50 ± 1.00 | 20.29 ± 1.71 | 0.049 |
| Sensation of urge to defecation | 24.64 ± 1.40 | 29.14 ± 2.35 | 0.089 |
| Maximally tolerable distension | 34.81 ± 1.65 | 39.11 ± 2.37 | 0.132 |
| IBS-SSS | 186.83 ± 8.42 |  |  |

*Note:* The data are presented as mean ± s.e. IBS-SSS: IBS Symptom Severity Scale; IBS-D: diarrhea-predominant IBS; HC: healthy control. \*: Numbers of subjects: IBS-D 26, HC 17; #: Numbers of subjects: IBS-D 28, HC 17.

### Increased melatonin expression was significantly negatively correlated with visceral sensitivity in IBS-D patients

Melatonin cell density was significantly higher in the colonic mucosa of IBS-D patients (94.88 ± 37.84/mm^2^) than in that of HC (73.61 ± 31.19/mm^2^) (*P* = 0.0318) **(Figure 1A, 1B, 1C)**. The arylalkylamine N-acetyltransferase (AANAT, a melatonin-synthesizing enzyme) cell density was also significantly increased in IBS-D patients (33.23 ± 24.34/mm^2^) compared to HC (24.50 ± 21.63/mm^2^) (*P* = 0.0214) **(Figure 1D, 1E, 1F)**.

**Figure 1.**
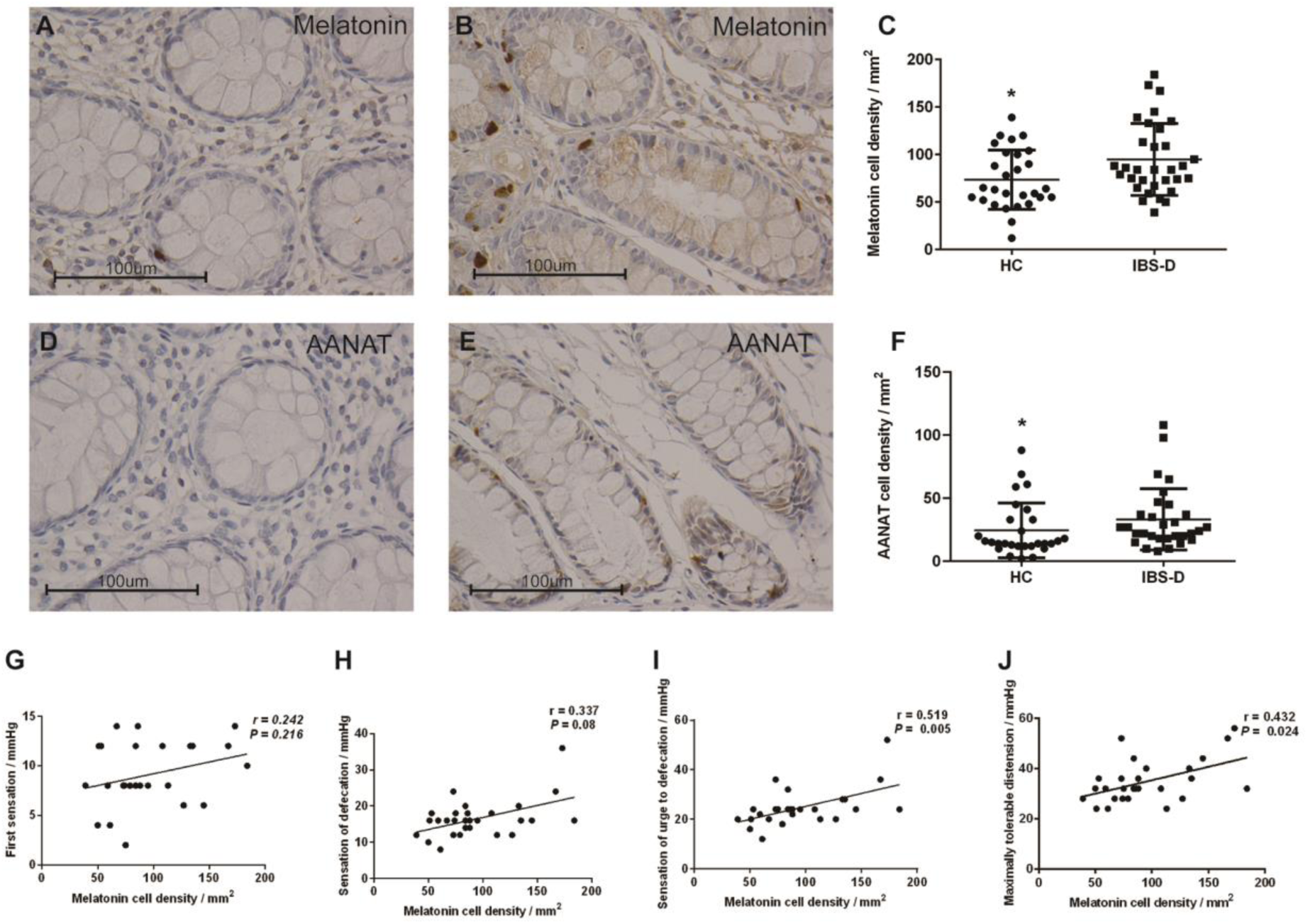
Increased melatonin expression was negatively associated with visceral hypersensitivity in IBS-D patients. Immunohistochemical staining of melatonin and AANAT cells in the colon of HC (*N* = 28) (**A, D**) and patients with IBS-D (*N* = 32) (**B, E**) at x 400 magnification. As shown in **C** and **F**, the densities of melatonin cells and AANAT cells were significantly higher in IBS-D patients than in healthy controls. The line in the scatter plots (**C, F**) indicates the mean value. Melatonin cell density was negatively associated with visceral hypersensitivity in IBS-D patients (**G, H, I, J**). * 0.01 < *P* ≤ 0.05. AANAT: arylalkylamine N-acetyltransferase; IBS-D: diarrhea-predominant IBS; HC: healthy controls.

To determine the role of melatonin in the GIT, it was necessary to determine whether IBS-D patients exhibited dysregulated melatonin expression (particularly mucosal melatonin expression), and this study was the first to evaluate the altered expression of mucosal melatonin in IBS patients. Melatonin synthesized in the GIT has been reported to be absorbed into the circulation and metabolized to 6-hydroxymelatonin sulfate (6-HMS) [17]. The 24-h urinary excretion of 6-HMS was also reported to be higher in the IBS-D patients, which supports our finding [18]. However, the results remain controversial. Another study reported the opposite results and found that the 24-h urinary excretion of 6-HMS was higher in the HC group [19]. Therefore, the alteration of gut melatonin may be a consequence rather than a cause according to the contradictory results regarding the melatonin metabolite 6-hydroxymelatonin sulfate in different studies.

To investigate the possible role of increased melatonin expression in IBS-D patients, we performed Spearman correlation analyses between melatonin level and visceral hypersensitivity in IBS-D patients. We found that the density of melatonin cells was positively correlated with sensation thresholds of the urge to defecate and maximal tolerable distension in IBS-D patients (**Figure 1G, 1H, 1I, 1J**), which supported the idea that increased melatonin levels may be a countermeasure for the development of IBS. No correlation was found between the density of melatonin cells and other parameters (psychological features and IBS Symptom Severity Scale (IBS-SSS) scores) **(Table 2)**.

**Table 2.** Correlations between colonic mucosal melatonin cell density with clinical parameters in human.

|  | Melatonin cell density [r (*P* values)] |  |
| --- | --- | --- |
|  | IBS-D | HC |
| Psychological feature |  |  |
| *Hospital anxiety scale | 0.207(0.310) | -0.372(0.141) |
| *Hospital depression scale | 0.007(0.972) | -0.414(0.099) |
| #Rectal distension (mmHg) |  |  |
| First sensation | 0.242(0.216) | -0.619 (0.018) |
| Sensation of defecation | 0.337(0.080) | -0.662 (0.010) |
| Sensation of urge to defecation | 0.519 (0.005) | -0.521(0.056) |
| Maximally tolerable distension | 0.432(0.024) | -0.373(0.140) |
| IBS-SSS scores | 0.068(0.720) |  |

*Note:* Spearman's correlation coefficients are shown as *P* values in parentheses. IBS-D: diarrhea-predominant IBS; HC: healthy control. IBS-SSS: IBS Symptom Severity Scale; \*: Numbers of subjects: IBS-D 26, HC 17; #Numbers of subjects: IBS-D 28, HC 17.

Low-grade inflammation has been confirmed in the colonic mucosa of IBS and mast cells are considered to play an essential role in mucosal inflammation [20]. In the present study, the mast cell count was significantly higher in the colonic mucosa of IBS-D patients (52.14 ± 29.86/mm^2^) than in that of HC (16.42 ± 12.01/mm^2^) (*P* = 0.000) (Figure 2A, 2B, 2C). Furthermore, mast cell counts were significantly negatively associated with the threshold of maximal tolerable distension in patients with IBS-D (Figure 2D, 2E, 2F, 2G). Numerous studies have suggested an increased mast cell count in the colons of patients with IBS [21]. In addition, an increased mast cell count and activation in the colons of patients with IBS may play an important role in the pathophysiology of IBS, especially in visceral hypersensitivity [3,22]. All of these findings are in line with our results Moreover, melatonin has been confirmed to inhibit the activation and proliferation of mast cells by in vitro and in vivo studies [23–25]. In our study, no significant differences were detected between the densities of melatonin cells and mast cells (r = 0.111, *P* = 0.567). Interestingly, when the mast cell counts were considered between one standard deviation above the mean density of the HC (28.43 / mm^2^) and nine standard deviations below the mean density of the HC (124.51 / mm^2^), the density of melatonin cells was significantly negatively correlated with the mast cell counts in patients with IBS-D (r = - 0.455, *P* = 0.038). In addition, accumulating evidence has demonstrated the protective effects of melatonin on the GIT, including immune enhancement, antioxidant activity, regulation of disturbed GI motility, visceral sensitivity reduction, and anxiolytic and antidepressant activity [8]. Above all, the elevated level of colonic mucosal melatonin in IBS-D patients may be based on an adaptive mechanism to modulate the dysfunction of the GIT.

**Figure 2.**
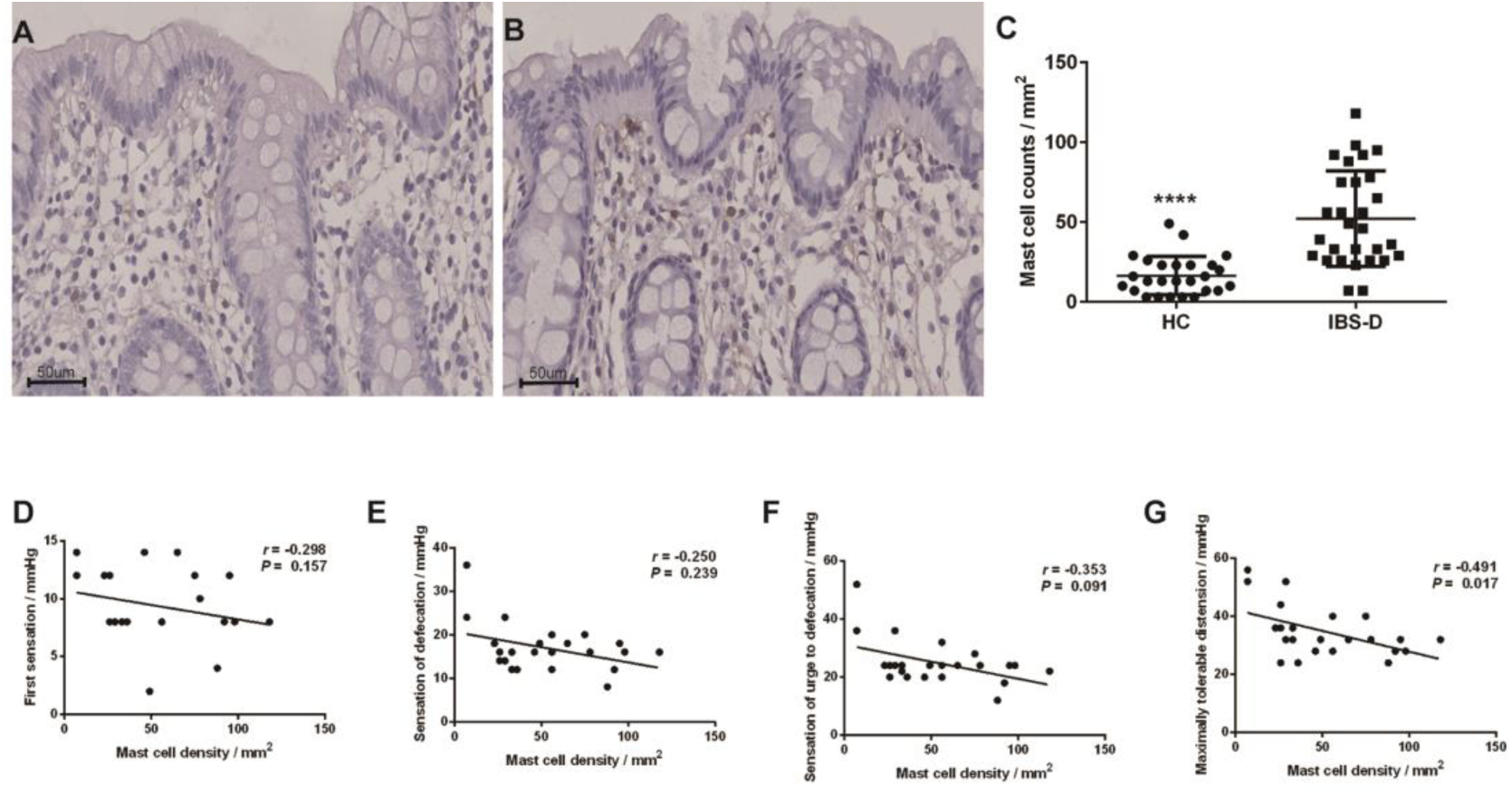
Increases in mast cells positively correlated with visceral hypersensitivity of IBS-D patients. Immunohistochemical staining of mast cells in the colon of HCs (N = 26) (A) and patients with IBS-D (N = 29) (B) at x 400 magnification. The densities of mast cells were significantly higher in IBS-D patients than in HC (C). Mast cell counts were positively correlated with visceral hypersensitivity in IBS-D patients (D, E, F, G). The line in the scatter plots (C) indicates the mean value. IBS-D: diarrhea-predominant IBS; HC: healthy controls. **** P ≤ 0.0001

### Transplantation of IBS-D patient fecal microbiota significantly elevated melatonin expression in germ-free (GF) rats

To further explore the correlations between gut dysbiosis and melatonin observed in patients with altered expression, we performed fecal microbiota transplantation (FMT) in male GF rats. We chose male GF rats for two reasons. First, estrogen in female rats has been confirmed to be involved in the regulation of motor and sensory function in the GIT and may confound the results [26]. Second, the gut microbiota has been suggested to be influenced by estrogen; thus, the composition of gut microbiota may be altered during the physiological cycle [27,28]. The density of melatonin cells in the GI group (GF rats receiving fecal specimens from one IBS-D patient) (57.83 ± 13.78 / mm^2^) was significantly higher than that in the GH group (GF rats receiving fecal specimens from one healthy control) (25 ± 10.46 / mm^2^) and GN group (GF rats without FMT) (20.2 ± 6.26 / mm^2^) **(Figure 3)**. The increased level of mucosal melatonin expression indicates that abnormal gut flora results in abnormal melatonin secretion in the colon via direct or indirect mechanisms.

**Figure 3.**
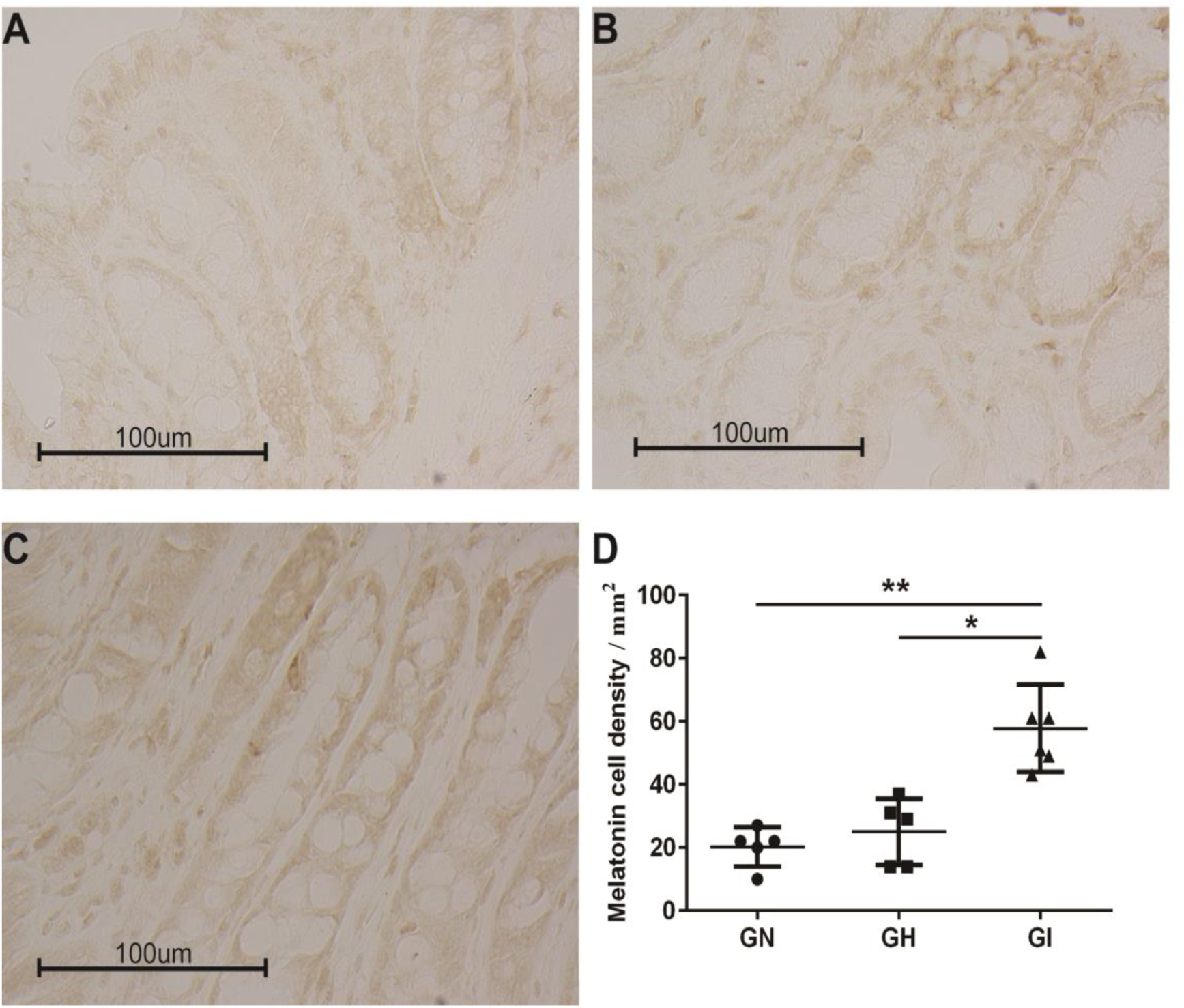
Melatonin expression in fecal microbiota transplantation (FMT) rats. Immunohistochemical staining of melatonin in the colon of the GN group (**A**), GH group (**B**), and GI group (**C**) at x 400 magnification. As shown in (**D**), the density of melatonin cells was significantly higher in the GI group than in the GH and GN groups. The line in the scatter plots indicates the mean value. * 0.01 < *P* ≤ 0.05, ** 0.001 < *P* ≤ 0.01. GI, Germ-free rats received fecal microbiota transplantation obtained from one IBS patient; GH, Germ-free rats received fecal microbiota transplantation obtained from one healthy control. GN, Germ-free without FMT.

The GF rats transplanted with the abnormal fecal microbiota from the IBS-D patient also exhibited visceral hypersensitivity. Abdominal withdrawal reflex (AWR) scores were significantly higher in GI rats than in GH rats regardless of the pressure applied (from 20 to 80 mmHg) (Figure S2), which is in accordance with a previous study in which fecal microbiota from a single IBS patient were transferred to GF rats [29]. In the colonic mucosa specimens, the density of mast cells was higher in GI rats (118.2 ± 64.02/mm^2^) than in GH rats (93.20 ± 32.90/mm^2^) (Figure S3); however, this difference was not significant.

### Gut dysbiosis was detected in IBS-D patients

Fecal samples from twenty-two IBS-D patients and eighteen HCs were collected for microbial analysis. The fecal microbiota richness in IBS-D patients was significantly increased compared with that in HCs based on the ACE (246.78 ± 84.07 versus 197.62 ± 48.88, respectively; *P* = 0.035) and Chao (243.16 ± 71.85 versus 198.36 ± 50.89, respectively; *P* = 0.032) richness indices. No significant differences in the Shannon (3.27 ± 0.60 versus 3.08 ± 0.43, *P* = 0.33) or Simpson (0.10 ± 0.089 versus 0.11 ± 0.058, *P* = 0.60) diversity indices were found between the two groups (Figure S4, Table S1). There were 636 operational taxonomic units (OTUs) in the IBS-D group and 524 OTUs in the HC group. In total, 463 OTUs coexisted in the two groups. The microbial community in the two groups could not be separated by principal coordinates analysis (PCoA) (Figure S5), which may be due to the heterogeneity within each group and the relatively small sample size. However, the partial least squares-discriminant analysis (PLS-DA) revealed separation according to groups (HC vs IBS-D) and was significantly different based on the cross-validation ANOVA (CV-ANOVA) (P = 0.000321217) **(Figure 4A)**. PLS-DA yielded a model with five components with a cumulated index (Q^2^ = 0.492) that measures the overall goodness of fit of the extremely high model explaining 98.1% (R^2^Y) of the variability between the HC and IBS-D groups. An analysis of similarity (ANOSIM) (R = 0.0659, *P* = 0.041, permutation number = 999) was also performed. A nonsignificant increase in *Firmicutes* and a decrease in *Bacteroidetes* were found in the IBS-D group in this study based on the community bar plot, which is in accordance with previous studies [30,31] **(Figure 4B)**. Moreover, the alteration in abundance of *Firmicutes* and *Bacteroidetes* in patients with IBS has been shown in many studies, but the results were controversial [32,33]. The reason for this discrepancy may be due to sample size, different DNA extraction methods and the various dietary habits of the recruited subjects [34]. The abundance of *Prevotella_9* was significantly increased in the IBS-D group compared to the HC group based on the Wilcoxon rank-sum test at the genus and species levels **(Figure 4C, 4D)**. *Prevotella* has been reported to be enriched in IBS-D patients and is suggested to be associated with a high risk of IBS-D [35]. The enrichment and reduction in the observed bacterial communities in IBS-D patients are displayed in a Manhattan plot **(Figure 4E)**. The percentage of community abundance at the genus level in the two groups is shown in Table S2.

**Figure 4.**
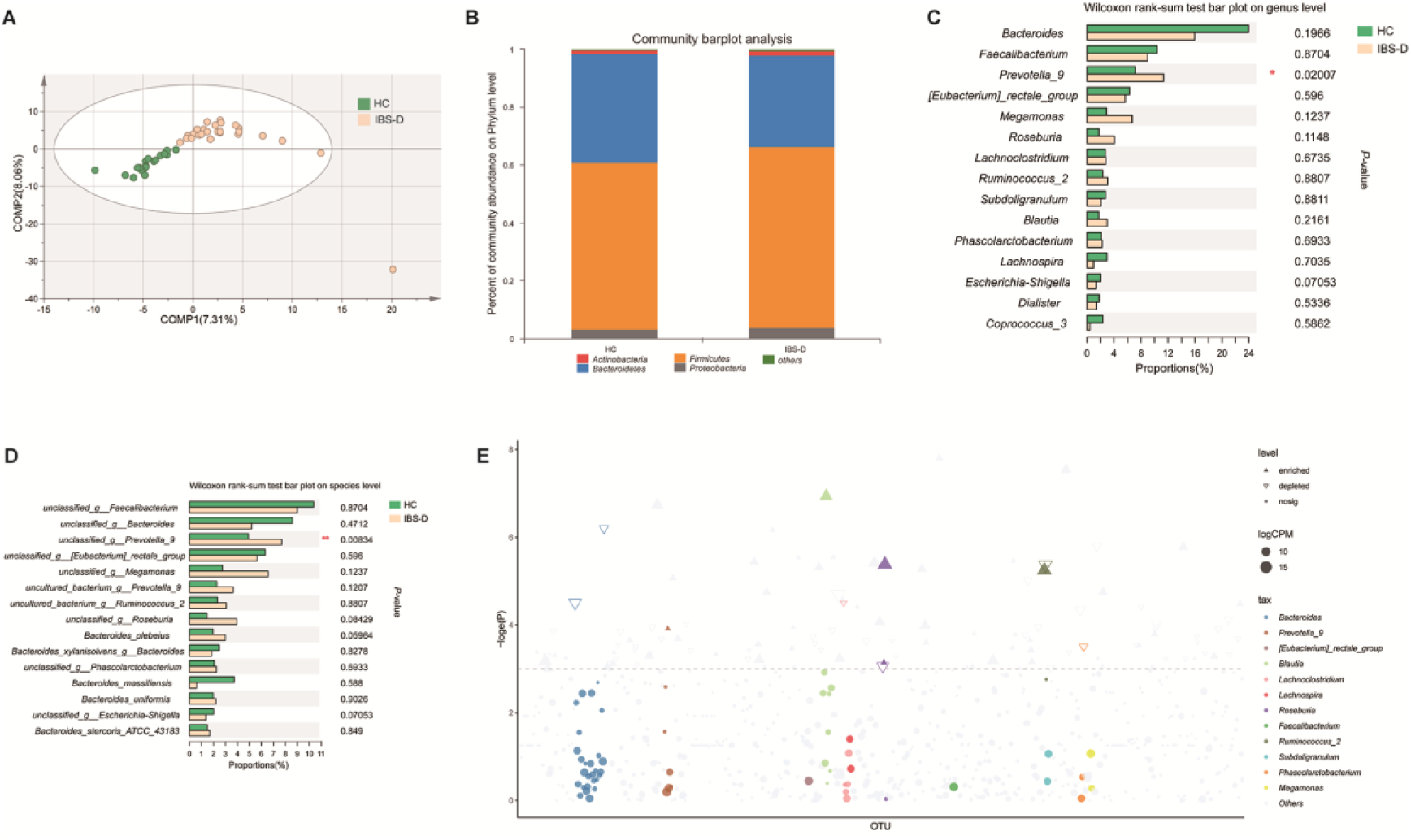
Gut dysbiosis in IBS-D patients. **A**. Partial least squares-discriminant analysis (PLS-DA) revealed separation in microbial composition between patients with IBS-D and HC. **B**. The abundance of *Firmicutes* was increased, and the abundance of *Bacteroidetes* was reduced in IBS-D patients compared with HC. **C, D**. Wilcoxon rank-sum test revealed that the abundance of *Prevotella_9* was significantly increased in IBS-D patients at the genus and species levels. **E**. Manhattan plots showing enriched and reduced OTUs in IBS-D patients. OTUs that are significantly enriched are depicted as full triangles. The dashed line indicates the threshold of significance (*P* = 0.05). The color and size of each dot represents the different taxonomic affiliation of the OTUs and relative abundance, respectively (genus level). * 0.01 < *P* ≤ 0.05. IBS-D: diarrhea-predominant IBS; HC: healthy controls.

Additionally, no significant difference was found in the amount of fecal SCFAs (short-chain fatty acids; the main fermentation products of microbiota) between the two groups (Figure S6), which may be due to the following reasons. First, the content of fecal butyrate depends on production by bacterial fermentation and absorption in the colon. Second, the relatively small sample size may be a factor. The multifactorial determinants of fecal SCFAs may contribute to the inconsistent results.

### Significant difference in gut microbiota in the GI and GH groups

The fecal microbiota richness in the GI group was remarkably higher than that in the GH group based on the ACE (137.24 ± 4.65 versus 94.26 ± 8.59, respectively; *P* < 0.000), Shannon (3.17 ± 0.12 versus 2.91 ± 0.21, respectively; *P* = 0.028), and Chao (137.61 ± 5.03 versus 94.06 ± 9.27, respectively; *P* < 0.000) richness indices. There were no significant differences in the Simpson (0.077 ± 0.012 versus 0.095 ± 0.043, *P* = 0.35) diversity index between the two groups (Figure S7, Table S3). The Venn diagram showed 153 OTUs in the GI group and 113 OTUs in the GH group. In total, 91 OTUs appeared simultaneously in the two groups. A significant difference in microbial composition was found between different groups by PCoA **(Figure 5A)**. The abundance of *Firmicutes* was significantly enriched, and the abundance of *Bacteroidetes* was significantly reduced in the GI group compared to the GH group based on a community bar plot **(Figure 5B)**.

**Figure 5.**
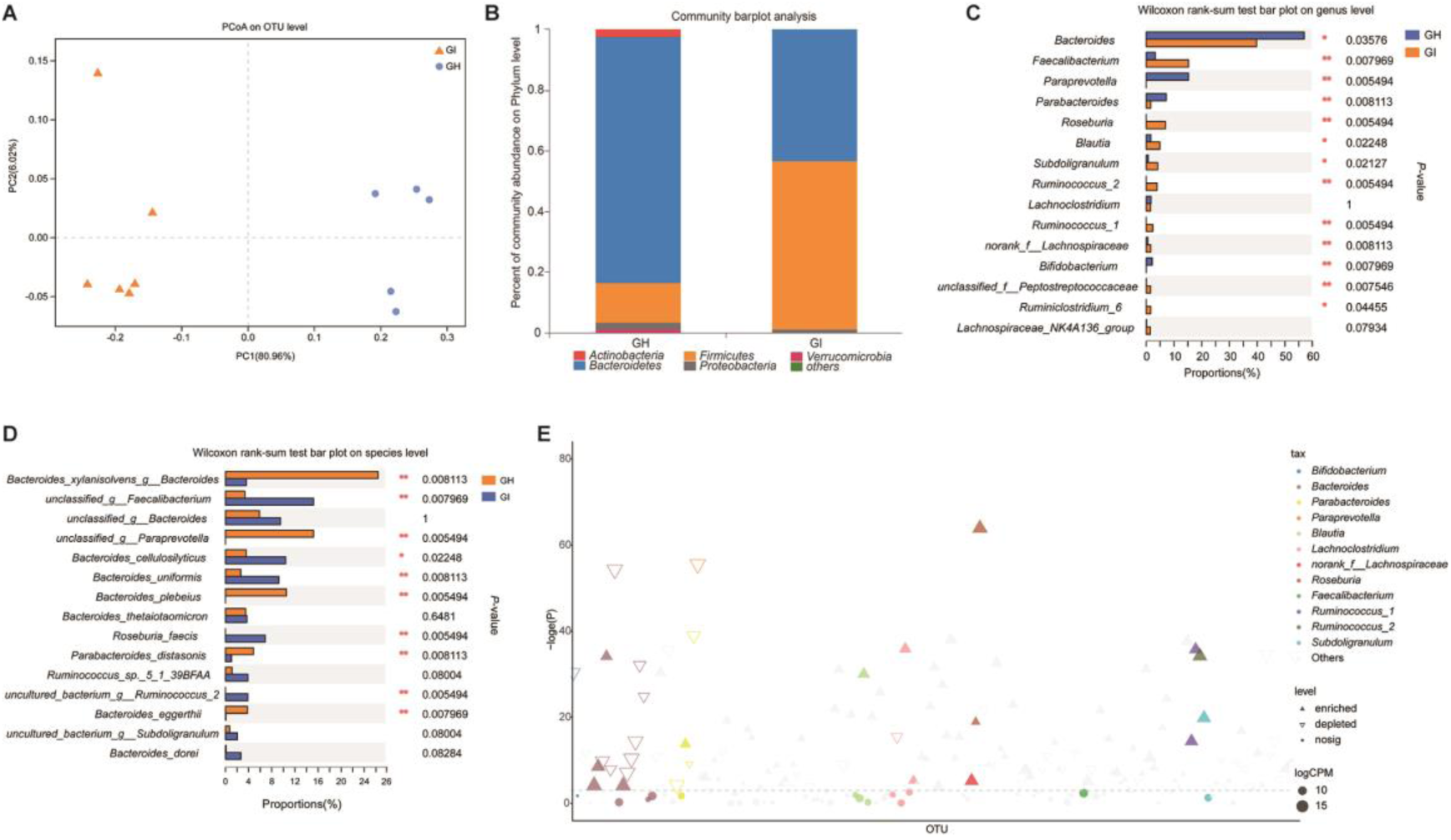
Significant differences in gut microbiota between the GI and GH groups. **A**. Principal coordinates analysis (PCoA) indicated significant differences in microbial composition between the GI group and GH group. **B**. The abundance of *Firmicutes* was increased, and the abundance of *Bacteroidetes* was reduced in the GI group based on a community bar plot. **C, D**. Wilcoxon rank-sum test revealed that the abundances of *Faecalibacterium, Roseburia*, and *Ruminococcus* were significantly increased and that the abundance of *Bacteroides* was significantly reduced in the GI group. **E**. Manhattan plots showing enriched and reduced OTUs in the GI group. OTUs that are significantly enriched are depicted as full triangles. The dashed line indicates the threshold of significance (*P* = 0.05). The color and size of each dot represent the different taxonomic affiliation of the OTUs and relative abundance, respectively (genus level). * 0.01 < *P* ≤ 0.05, ** 0.001 < *P* ≤ 0.01. GH, Germ-free rats received fecal microbiota transplantation obtained from one healthy control. GI, Germ-free rats received fecal microbiota transplantation obtained from one IBS patient.

The abundances of *Faecalibacterium, Roseburia*, and *Ruminococcus* were significantly enriched, and the abundance of Bacteroides was significantly reduced in the GI group based on the Wilcoxon rank-sum test at the genus and species levels **(Figure 5C, 5D)**. The enrichment and reduction in the observed bacterial communities are displayed in a Manhattan plot **(Figure 5E)**. The percentages of community abundances at the genus level in FMT rats are shown in Table S4.

The amounts of fecal formate, butyrate, valerate, and isovalerate were remarkably increased, and the amount of fecal propionate was remarkably reduced in the GI group compared with the GH group (Figure S8).

### Some butyrate-producing *Clostridium* cluster *XIVa* bacteria species were positively associated with colonic mucosal melatonin expression

As shown in the heat maps of Spearman rank correlation coefficients in humans, *Roseburia* was significantly positively correlated with melatonin and AANAT expression. *Lachnospira* and *Parasutterella* were significantly positively associated with AANAT expression **(Figure 6A)**. Additionally, *Roseburia* and *Lachnospira* were significantly positively related to fecal butyrate levels. The genus *Roseburia*, which belongs to *Clostridium* cluster XIVa, consists of *Roseburia intestinalis, R. hominis, R. inulinivorans, R. faecis* and *R. cecicola* [36]. The ability of *Roseburia* species to penetrate the mucus layer enables them to interact with gut epithelial cell surfaces and exert physiological effects [37]. As SCFA producers, *Lachnospira* species also belong to *Clostridium* cluster XIVa [38,39]. On the one hand, *Clostridium* cluster XIVa species are known to be butyrate-producing bacteria and play a potential role in the regulation of gut motility and innate immunity (mainly anti-inflammatory effects) [36, 40]. Butyrate has been confirmed to stimulate the expression of colonic serotonin (a precursor of melatonin) in both in vivo and in vitro studies [41,42]. Moreover, butyrate has been reported to promote melatonin expression in an intestinal epithelial cell line (Caco-2 cells) [43], indicating that *Clostridium* cluster XIVa species may promote melatonin expression via butyrate. On the other hand, melatonin facilitates goblet proliferation and MUC2 expression [44]. As mucin-adherent bacteria, *Clostridium* cluster XIVa species compose 60% of mucosal microbiota, and stimulating mucus production may lead to a higher abundance of mucin-adherent bacteria [37]. Therefore, we postulate that melatonin could increase the abundance of *Roseburia* and *Lachnospira* via the promotion of mucus secretion, in turn stimulating melatonin expression and constituting a positive feedback loop.

**Figure 6.**
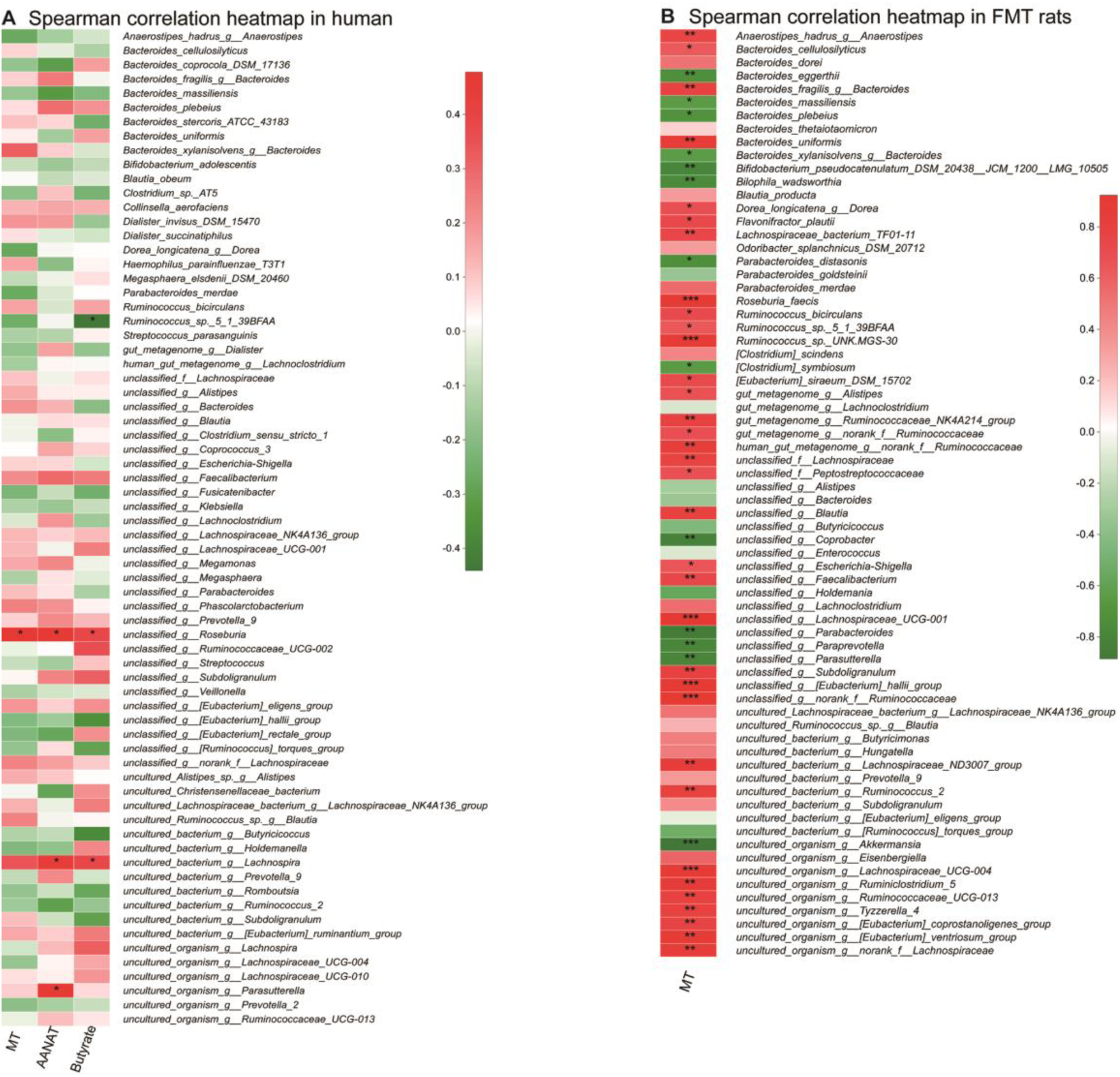
Relationship between melatonin and gut microbiota in humans. **A**. Heat maps of Spearman’s rank correlation coefficients between human colonic mucosal melatonin and AANAT expression, SCFAs and gut microbiota at the species level. *Roseburia* was significantly positively correlated with melatonin and AANAT expression. *Lachnospira* and *Parasutterella* were significantly positively associated with AANAT expression. *Roseburia* and *Lachnospira* were significantly positively related to fecal butyrate. **B**. The relationship between melatonin and gut microbiota in FMT rats. Heat maps of Spearman’s rank correlation coefficients between FMT rat colonic mucosal melatonin expression and gut microbiota at the species level. *Roseburia, Lachnospiraceae, Blautia* and *Ruminiclostridium* species, which belong to *Clostridium* cluster XIVa, were significantly positively related to melatonin expression. * 0.01 < *P* ≤ 0.05, ** 0.001 < *P* ≤ 0.01, *** *P* ≤ 0.001.

In view of the correlation analyses in GF rats, most of the bacteria that positively correlated with melatonin expression belonged to *Clostridium* cluster XIVa such as *Roseburia, Lachnospiraceae, Blautia* and *Ruminiclostridium* species **(Figure 6B)**, which is consistent with the results in humans. Furthermore, the changes in the abundance of butyrate-producing *Clostridium* cluster XIVa species (such as *Roseburia*) in IBS patients were not consistent among different studies [45,46] indicating that these changes may also be a consequence rather than a cause of the disease.

### Sodium butyrate promotes melatonin secretion in BON-1 cells

To confirm the effect of butyrate, one of the main metabolites of butyrate-producing *Clostridium* cluster XIVa bacteria species, we used sodium butyrate to treat BON-1 cells and detected the melatonin concentration and AANAT expression in the treated cells using ELISA and western blots, respectively. After treatment with sodium butyrate for 24 h, the melatonin level was significantly increased in a dose-dependent manner in BON-1 cells **(Figure 7)**. The 10 mM sodium butyrate treatment group showed a significant increase in melatonin secretion compared with the untreated control group and the 2.5 mM sodium butyrate-treatment group. The effect of butyrate on AANAT was investigated by western blot assays. Treatment with 5 and 10 mM sodium butyrate promoted AANAT expression in BON-1 cells, but the difference was not significant. Therefore, butyrate could increase melatonin expression in a dose-dependent manner according to our research, which was consistent with a previous study [43]. Moreover, many studies have suggested that butyrate can promote tight junction-mediated barrier integrity and exert anti-inflammatory functions. Considering the protective effects of melatonin, the butyrate-melatonin link may be a protective mechanism of butyrate in IBS.

**Figure 7.**
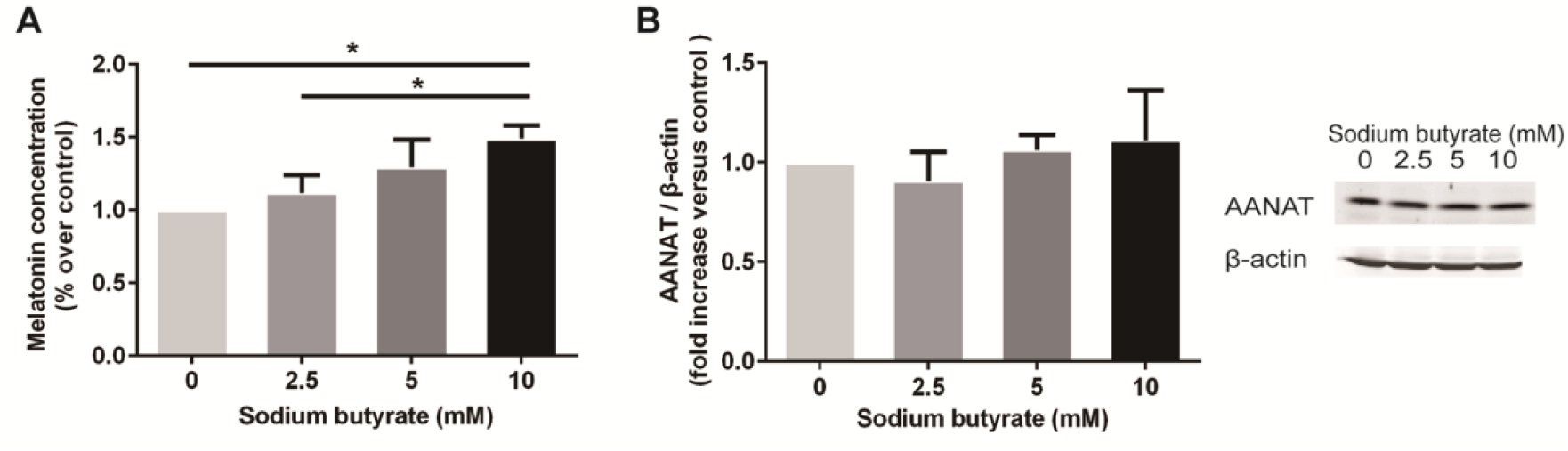
Promotion of butyrate on melatonin and AANAT expression in BON-1 cells. Levels of melatonin in the supernatants were detected using ELISA. AANAT expression was detected with western blot assays. **A**. Treatment with sodium butyrate significantly increased melatonin secretion in a dose-dependent manner. **B**. Relative AANAT quantification by western blot analysis of protein extract of control or sodium butyrate-treated BON-1 cells. The results are presented as the mean ± SEM. * 0.01 < *P* ≤ 0.05. AANAT: arylalkylamine N-acetyltransferase

There were several limitations in this study. First, the present study involved only patients with IBS-D, and additional studies are needed to evaluate the alteration of melatonin in patients with other IBS subtypes. Second, cause and effect inference cannot be made based on our research. Further studies are needed to explore the potential mechanisms underlying the association between gut microbiota and mucosal melatonin.

In conclusion, our study is the first to demonstrate that melatonin expression is increased in the colonic mucosa of IBS-D patients. Increased melatonin expression may be an adaptive protective mechanism to reduce visceral hypersensitivity. Notably, the study assessed the relationships between gut microbiota and melatonin expression and suggested that different microbiota structures may contribute to different colonic mucosal melatonin levels. Moreover, some butyrate-producing *Clostridium* cluster XIVa species, such as *Roseburia* and *Lachnospira*, could promote melatonin expression via butyrate. These preliminary findings provide some clues for the involvement of the dysregulated gut hormone/gut microbiota axis in the elusive pathophysiology of IBS and in the search for new targeted approaches in the treatment of IBS.

## Methods and materials

### Subjects

Patients with IBS-D (aged 18–65 years) were enrolled from the gastroenterology clinic of Peking University Third Hospital between March 2015 and January 2016 following the Rome III criteria [47]. Healthy volunteers without any symptoms (aged 18-65 years) were recruited by an Internet advertisement. The inclusion criteria were as follows: 1) normal colonoscopy and 2) no abnormalities in routine blood or stool exams or in liver or kidney function. The exclusion criteria were as follows: 1) organic gastrointestinal or systemic diseases, such as inflammatory bowel diseases or diabetes mellitus; 2) current infectious diseases of the respiratory, digestive, or urinary system; 3) a history of abdominal surgery; 4) history of antibiotic or probiotic use during the previous 4 weeks; 5) continuous laxative or antidiarrheal use for more than 3 days; 6) lactating or pregnant women; and 7) mental illness. Symptom severity was measured with the IBS-SSS. The severity of anxiety and depression symptoms was evaluated with the Hospital Anxiety and Depression Scale (HADS). Eating habits were investigated with a three-day dietary record sheet. Fecal samples were collected for sequencing on the Illumina MiSeq platform. Visceral sensitivity was assessed by rectal barostat, as previously described in detail [48]; sensory thresholds for first sensation, sensation of defecation, sensation of urge to defecate and maximal tolerable distension were recorded as the lowest distention pressures (mmHg). All participants underwent colonoscopy. Sigmoid colonic mucosal biopsies were obtained during colonoscopy. The study was approved by the Ethics Committee of Peking University Health Science Center (RB00001052—14091). Written informed consent was obtained from all participants.

### Immunohistochemistry

Sections were cut with a Leica microtome at a thickness of 4 μm. The sections were deparaffinized by following routine procedures. Rabbit anti-melatonin (abx100179, 1:800, Abbexa, Cambridge, UK), rabbit anti - AANAT (orb5712, 1:800, Biorbyt, Wuhan, China), and anti-mast cell tryptase (ab2378, 1:3000, Abcam, Cambridge, UK) primary antibodies were used to detect melatonin, AANAT, and mast cells, respectively. A horseradish peroxidase/3,3’ - diaminobenzidine detection system (PV-6000, PV-6001, Zhongshan Golden Bridge, Beijing, China) was used for visualization.

For the quantification of melatonin and AANAT expression, ten fields were randomly selected at a magnification of x400 from each slide under a Leica DM 2500 microscope (LEICA Microsystems, Germany), captured using a Nikon DS-Vi1 camera (Nikon Instruments, China), and exported by the Nikon NIS-Elements imaging software. The number of immunoreactive cells was counted in every field and the area of every field was calculated using Image-Pro Plus 6.0 software (Media Cybernetics, Inc., USA). To quantify mast cells, the NanoZoomer-SQ digital slide scanner (Hamamatsu Photonics, Japan) was used for whole slide scanning, and three randomly selected fields at a magnification of x400 were exported by NDP view2 viewing software (Hamamatsu Photonics, Japan). The density of immunoreactive cells was expressed as the average number of cells per square millimeter of mucosal epithelium.

### DNA extraction of fecal microbiota

Microbial DNA was extracted from fecal samples with an E.Z.N.A.® Soil DNA Kit (Omega Bio-Tek, Norcross, GA, USA). Two primers [338F regions (V3 to V4) of the bacterial 16S ribosomal RNA gene. Polymerase chain reaction was performed with the following program: 3 min of denaturation at 95C; 27 cycles of 95°C for 30 s, 55°C for 30 s, and 72°C for 45 s; and a final extension at 72°C for 10 min. Purified amplicons were pooled in equimolar amounts, an d paired-end sequencing (2×300) was performed on the Illumina MiSeq platform (Illumina, San Diego, CA, USA).

### Processing of sequencing data

Trimmomatic was used to demultiplex and quality-filter raw FASTQ files and raw FASTQ files were merged by fast length adjustment of short reads (FLASH). OTU clustering was performed by UPRISE (version 7.0 http://drive5.com/uparse/). Chimeric sequences were identified and removed by UCHIME. The taxonomy of each 16S rRNA gene sequence analysis was performed using the RDP Classifier algorithm against the Silva (SSU128) 16S rRNA database with confidence threshold of 70%.

### Quantification of fecal SCFAs

Ultra-performance liquid chromatography coupled with tandem mass spectrometry (UPLC-MS/MS) was used to determine the levels of SCFAs (formate, acetate, propionate, butyrate, isobutyrate, valerate, and isovalerate acetate) in feces as previously described [49].

### Animal experiments

Male germ-free (GF) Sprague-Dawley (SD) rats were obtained from the Department of Laboratory Animal Science at Peking University Health Science Center. GF rats were divided into three groups and housed in separated sterilized isolators. Different groups of GF rats were maintained in different sterile bubbles, and every GF rat was housed in a separate cage to avoid cage effects. A total of 16 GF rats (6 in the GI group, 5 in the GH group, and 5 in the GN group) were used in this study. At six weeks of age, GF rats in different groups were subjected to fecal microbiota transplantation [obtained from one IBS-D patient (GI group) and one healthy control (GH group)] separately by gavage. Rats in the same group received fecal microbiota from the same sample, and transplantation was performed only once. Two weeks after FMT, fecal samples were collected from the rats for further microbiota analyses. Colorectal distension (CRD) tests were performed 24 days after FMT as described previously [48]. Then, GF rats were sacrificed with anesthesia. Colon tissue was fixed in 10% buffered formalin for immunohistochemistry. All protocols were approved by the Laboratory Animal Welfare Ethics branch of the Biomedical Ethics Committee of Peking University (LA2016230).

### Cell culture

BON-1 cells were purchased from Shanghai Shun Ran Biotechnology Co., Ltd. and cultured in DMEM (SH30022.01, HyClone, Utah, USA) supplemented with 10% FBS (900-108, Gemini, California, USA) and 1% penicillin/streptomycin (15140122, Gibco, Massachusetts, USA) at 37 °C in a humidified atmosphere of 5% CO2. Cells were subcultured every 2 days.

### Melatonin measurements

Cells were seeded in 60-mm dishes, allowed to attach and treated with sodium butyrate (303410, Sigma-Aldrich, USA) at concentrations ranging from 0 to 10 mM for 24 h. Then, supernatants were harvested and melatonin levels were determined with an ELISA kit (E-EL-H2O16c, Elabscience, Wuhan, China). Readings were normalized to total protein content, which was detected using a BCA assay. The data compiled from different treatment groups were expressed as melatonin concentrations normalized to the untreated control group.

### Western blot analysis

Proteins from BON-1 cells were extracted with RIPA extraction buffer mixed with a protease inhibitor cocktail. Equal amounts of protein (30 µg) were electrophoresed in 12% sodium dodecyl sulfate polyacrylamide gels and transferred onto nitrocellulose membranes under a constant current of 250 mA for 90 min. The membranes were incubated with rabbit anti-AANAT antibody (1:1,000, ab3505, Abcam, Cambridge, UK) overnight at 4°C. β-actin was chosen as the internal reference protein. The primary antibodies were detected using anti-rabbit or mouse secondary antibodies and were visualized with an infrared imaging system (LI-COR Biosciences, Lincoln, USA). The band intensity relative to that of β-actin was calculated, and the results were expressed as fold change from control values.

### Statistical analysis

The mean number of melatonin and AANAT immunoreactive cells per square millimeter of mucosal epithelium was compared using the nonparametric Mann-Whitney test (between IBS-D patients and HC) and Kruskal-Wallis nonparametric test with posttest (among different GF rats). The differences in age and BMI were analyzed by independent samples t-test. The ^2^ test or Fisher’s exact test was used to detect sex differences between IBS-D patients and HC. Correlations between melatonin and clinical and experimental parameters were explored using Spearman’s rank correlation analysis. The statistical analyses were performed using SPSS software version 24.0 (SPSS Inc., Chicago, IL, USA). Alpha diversity based on the OTU table was analyzed with the abundance-based coverage estimator (ACE), the Chao1 estimator (Chao), and the Shannon and Simpson diversity indices in Mothur (version v.1.30.1). β-Diversity was calculated using UniFrac distance, and PCoA was performed in R (version 3.3.1) to visualize the difference in community composition among different groups. PLS-DA and CV-ANOVA were performed using SIMCA (version 14.1.0). The associations among the expression levels of melatonin, AANAT, SCFAs (acetate, propionate, butyrate) and bacterial taxa were visualized and identified by heat maps of Spearman rank correlation coefficients using the pheatmap package in R (version 3.3.1). One-way analysis of variance (ANOVA) with post hoc testing was conducted using SPSS to determine differences in melatonin secretion according to ELISA among different treatment groups.

## Data Availability

The raw sequence data have been uploaded to the Genome Sequence Archive in BIG Data Center under accession number CRA001604 and are publicly accessible at http://bigd.big.ac.cn/gsa.

## Data availability

The raw sequence data have been uploaded to the Genome Sequence Archive [50] in BIG Data Center [51] under accession number CRA001604 and are publicly accessible at http://bigd.big.ac.cn/gsa.

## Author contributions

DL and WB designed the study; ZS, lZ, WH, ZL, SQ and WB recruited the IBS-D patients and HC. WB, ZS, lZ, WH, ZL, ZJ and SQ acquired data; WB drafted the initial manuscript; WB, ZS and HM analyzed and interpreted the data; HM and PF provided technical support; and DL revised the manuscript critically. All authors approved the final version of the manuscript.

## Competing interests

The authors have declared no competing interests.

## Acknowledgments

This study was supported by the National Natural Science Foundation of China (81670491) and the Capital Health Research and Development of Special Program (2016-2-4093). The authors are grateful to all subjects participating in this study. The authors appreciate the technical support of the Medical Research Center of Peking University Third Hospital and the Clinical Stem Cell Center of Peking University Third Hospital. The authors appreciate the support of the Department of Laboratory Animal Science, Peking University Health Science Center with the animal experiments. The authors appreciate the assistance of the Institute of Acupuncture and Moxibustion China Academy of Chinese Medical Sciences for the CRD tests in rats. The authors appreciate the technical assistance of Professor Ence Yang (Institute of Systems Biomedicine, School of Basic Medical Sciences, Peking University) and Professor Xiaoli Wang (School of Public Health, Peking University).

